# Survey of vitamin D intake in the general population in the Democratic Republic of Congo: Multicentre cross-sectional study

**DOI:** 10.1101/2024.04.24.24306262

**Authors:** Dieudonne Masemo Bihehe, Jacques Musung Mbaz, Bertin Isaga, Willy David Shamputi, Jean Luhaga Mulume, Jean Missel Selemani, Jean Mukonkole Mbo, Ahadi Birindwa Bwihangane, Eric Etale, Paul Tshonda Ngongo, Roland Cibenda Lwandiko, Philippe Bianga Katchunga

## Abstract

**Aim:** Data on vitamin D intake are not available for the Democratic Republic of Congo. The aim of this study was to determine the level of sun exposure and dietary intake of vitamin D in the general adult Congolese population.

**Methods:** Between January and March 2024, 14,750 respondents (age= 31.3±12.5 years, 53.5% male) completed a questionnaire on sun exposure and dietary vitamin D intake. The association between satisfactory vitamin D intake ≥ 15 µg/day and hypothesised determinants was modelled in a multiple logistic regression.

**Results:** Of the 14,750 participants, 1,238 (8.4%) had very fair skin, 4,601 (31.2%) had less than 15 minutes’ exposure to the sun, 5,281 (35.8%) had only their head exposed to the sun and 2,509 (17.0%) had a vitamin D test. Fish was the main source of vitamin D intake [3.0 (1.0-7.0) µg/100 gr/day]. Finally, satisfactory vitamin D intake was significantly more frequent in participants aged < 40 years (adjusted OR= 2.45; p=0.0007), in those with a low socio-economic level (adjusted OR= 1.23; p=0.01) and in one of the towns studied (adjusted OR= 1.64; p=0.0001).

**Conclusion:** A low level of sun exposure and dietary intake of vitamin D is observed in the general Congolese population. Vitamin D intake is influenced by age and socio-economic status. The introduction of a vitamin D programme integrated into funded programmes (TB, HIV) would make it possible, at a lower cost, to combat vitamin D deficiency by raising awareness, screening and treating the general Congolese population.

## I. Introduction

Vitamin D (Vit D) is an essential micronutrient with local and systemic effects. In fact, it plays an essential role in regulating blood calcium and phosphate levels and bone homeostasis [1]. Vit D also has a protective effect against infections, autoimmune diseases, cancers and cardiovascular diseases, including arterial hypertension, and metabolic diseases such as insulin resistance and type 2 diabetes mellitus [1–3]. As a result, Vit D intake remains essential for humans, hence the ever-growing scientific interest in this vitamin.

The sources of Vit D vary from region to region [4]. Endogenous intake is obtained from sunlight, which enables it to be synthesised in the skin, while exogenous intake comes from the diet and/or supplements [4].

But despite all these alternatives, Vit D deficiency is currently a global public health problem, affecting more than half the world’s population [5].

Vit D deficiency varies according to ethnicity, geographical location, exposure to sunlight, age, obesity and cultural and dietary habits [4]. More specifically, numerous dietary surveys show a very high level of dietary intake, well below the recommendations of scientific societies [6].

Sub-Saharan Africa (SSA) is the region with the longest period of sunshine in the world. Despite this, the prevalence of Vit D deficiency is very high and similar to that in other regions [7,8]. It is likely that certain cultural habits are not conducive to adequate sunlight. In addition, there are few natural food sources rich in Vit D in this region [8]. In addition, the low socio-economic level certainly limits access to these Vit D-rich food sources for a large proportion of the population [8]. Unfortunately, data on Vit D intakes in this region are scarce and unrepresentative.

Singularly, in the Democratic Republic of Congo (DRC), studies on Vit D are very rare. A few studies show a very high prevalence of Vit D deficiency in specific groups: asthmatics (95.2%) [9], pre-eclampsia patients (80.0%) [10] and diabetics (68.5%) [2]. However, to our knowledge, no study has investigated the reasons for this deficiency in the general population.

However, knowledge of the reasons for inadequate Vit D intake in the general population could lead to recommendations for improving the overall health of the population.

The aim of this study was therefore to determine the reasons of inadequate Vit D intake in a number of towns in the Democratic Republic of Congo.

## II. Participants and method

### II.1 Subjects surveyed

This study was cross-sectional and multicentre. It took place between 08 January 2024 and 10 March 2024 respectively in the city of Bukavu (pop=1,000,000) in the province of Sud-Kivu, in the city of Kindu (pop= 450,000) in the province of Maniema and in the city of Lubumbashi (pop= 1,200,000) in the province of Haut-Katanga.

A target population of 1% of adult subjects aged 18 and over, i.e. a sample of 13,300 subjects, was expected.

Respondents were found in their homes or in public places (universities, hospitals, churches, health centres, markets, workplaces, online questionnaires).

Informed verbal consent was obtained from each study participant. Data were collected anonymously and confidentially. The privacy and personality of the participants were protected in accordance with the Declaration of Helsinki.

The protocol for this study was accepted by the ethics committee of the official University of Bukavu. (UOB/CEM/013/2023).

### II.2 Data collection

Teams of interviewers made up of doctors and medical students from the Evangelical University in Africa (UEA), the official University of Bukavu (UOB), the University of Kindu (UNIKI) and the University of Lubumbashi (UNILU) were trained to collect data using a multiple-choice questionnaire in French or Swahili (the local language). The questionnaire contained the following information: demographic parameters (age, sex), socio-economic parameters (level of education, occupation, town of residence), skin colour, exposure to the sun (duration, part of body exposed to the sun), vitamin D screening and blood calcium levels, weekly consumption of Vit D-rich foods available in the study region (fresh or frozen fish, tinned fish, eggs, foods containing eggs (cakes, galettes, cake), meat/sausages, yoghurt/cream, cheese, green vegetables, bread, fruit and chocolate, vitamin D-containing medication taken. This questionnaire was adapted from the self-administered questionnaire proposed by Grados F at al [11].

### II.3 Operational definitions

In this study, we standardised Vit D intake by estimating, for each participant, the average Vit D content (µg/100g) provided by the food consumed, based on the Ciqual® nutritional composition tables proposed by the French National Agency for Food, Environmental and Occupational Health and Safety. (Table 1).

**Table 1.**
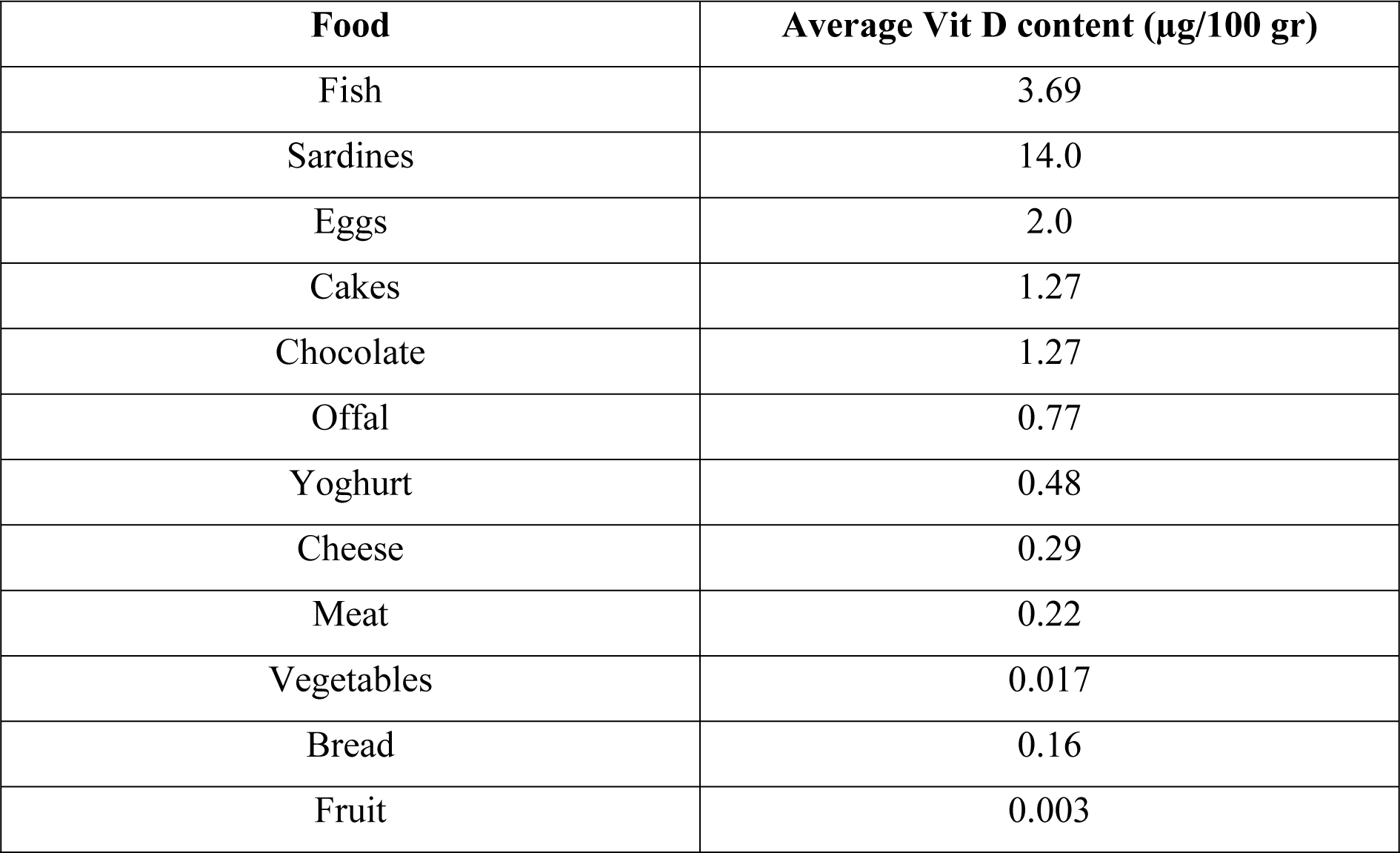
Average Vit D content of foods (ciqual)

Daily intake was considered satisfactory in adults when it was ≥ 15 µg/day.

Also, in this study, participants with a high socio-economic level were those with a university degree and a profession.

### II.4 Statistical analysis

The data from this study are presented in the form of means (± standard deviation), median (interquartile range) or frequency, as appropriate.

The Anova test was used to compare several means and the Chi² test to compare proportions. The association between demographic and socio-economic parameters and Vit D intake judged to be satisfactory was modelled using multiple logistic regression.

A probability of 0.05 or less was considered statistically significant. To do this, we used MedCalc® version 18.11 software.

## III. Results

### III.1 General characteristics of the study population

Table 2 illustrates the general characteristics of the population studied. A total of 14,750 subjects took part in this nutritional survey, 6,494 (44.0%) in the city of Lubumbashi, 6,347 (43.0%) in the city of Bukavu and 1,909 (12.9%) in the city of Kindu. The average age was 31.3±12.5 years.

**Table 2.** General characteristics of the study population.

|  | **Population**<br>**studied** | **Men** | **Women** | **p** |
| --- | --- | --- | --- | --- |
| **Cities** |  |  |  |  |
| Lubumbashi, n (%) | 6,494 (44.0) | 3,581 (55.1) | 2,913 (44.9) | <0.0001 |
| Bukavu, n (%) | 6,347 (43.0) | 3,049 (48.0) | 3,298 (52.0) | <0.0001 |
| Kindu, n (%) | 1,909 (12.9) | 1,254 (65.7) | 655 (43.3) | <0.0001 |
| Total | 14,750 (100) | 7,884 (53.5) | 6,866 (46.5) | <0.0001 |
| **Age (years), avg±SD** | 313±12.5 | 31.5±11.8 | 31.1±13.2 | 0.02 |
| <40 years, n (%) | 11,796 (80.0) | 6,297 (79.9) | 5,499 (80.1) | 0.76 |
| 40-59 years, n (%) | 2,172 (14.7) | 1,252 (15.9) | 920 (13.4) | <0.0001 |
| $\geq 60$ years, n (%) | 775 (5.3) | 330 (4.2) | 445 (6.5) | <0.0001 |
| **Level of education** |  |  |  |  |
| Illiterate | 902 (6.1) | 402 (5.1) | 500 (7.3) | <0.0001 |
| Primary | 1,121 (7.6) | 489 (6.2) | 632 (9.2) | <0.0001 |
| Secondary | 5,085 (34.5) | 2,523 (32.0) | 2,562 (37.3) | <0.0001 |
| University | 7,642 (51.8) | 4,470 (56.7) | 3,172 (46.2) | <0.0001 |
| **Profession** |  |  |  |  |
| Without | 5,936 (40.3) | 2,885 (36.6) | 3,051 (44.4) | <0.0001 |
| With | 8,811 (59.7) | 4,997 (63.4) | 3,814 (55.6) | <0.0001 |
| **High socio-economic level** | 4,689 (31.8) | 2,855 (36.2) | 1,834 (26.7) | <0.0001 |
| **Vit D3 dosage** | 2,509 (17.0) | 1,196 (15.2) | 1,313 (19.1) | <0.0001 |
| **Vit D supplements** | 3,593 (24.4) | 1,895 (24.0) | 1,698 (24.7) | 0.32 |

Still in the whole group, 7,642 (51.8%) were university graduates, 8,811 (59.7%) had a profession and 4,689 (31.8%) had a high socio-economic level.

### III.2 Skin type and level of sun exposure

Table 3 shows the skin type and level of sun exposure of the population studied. Of the 14,750 participants, only 1,238 (8.4%) had very fair skin, 769 (11.2%) women vs. 469 (5.9%) men (p<0.0001).

**Table 3.**
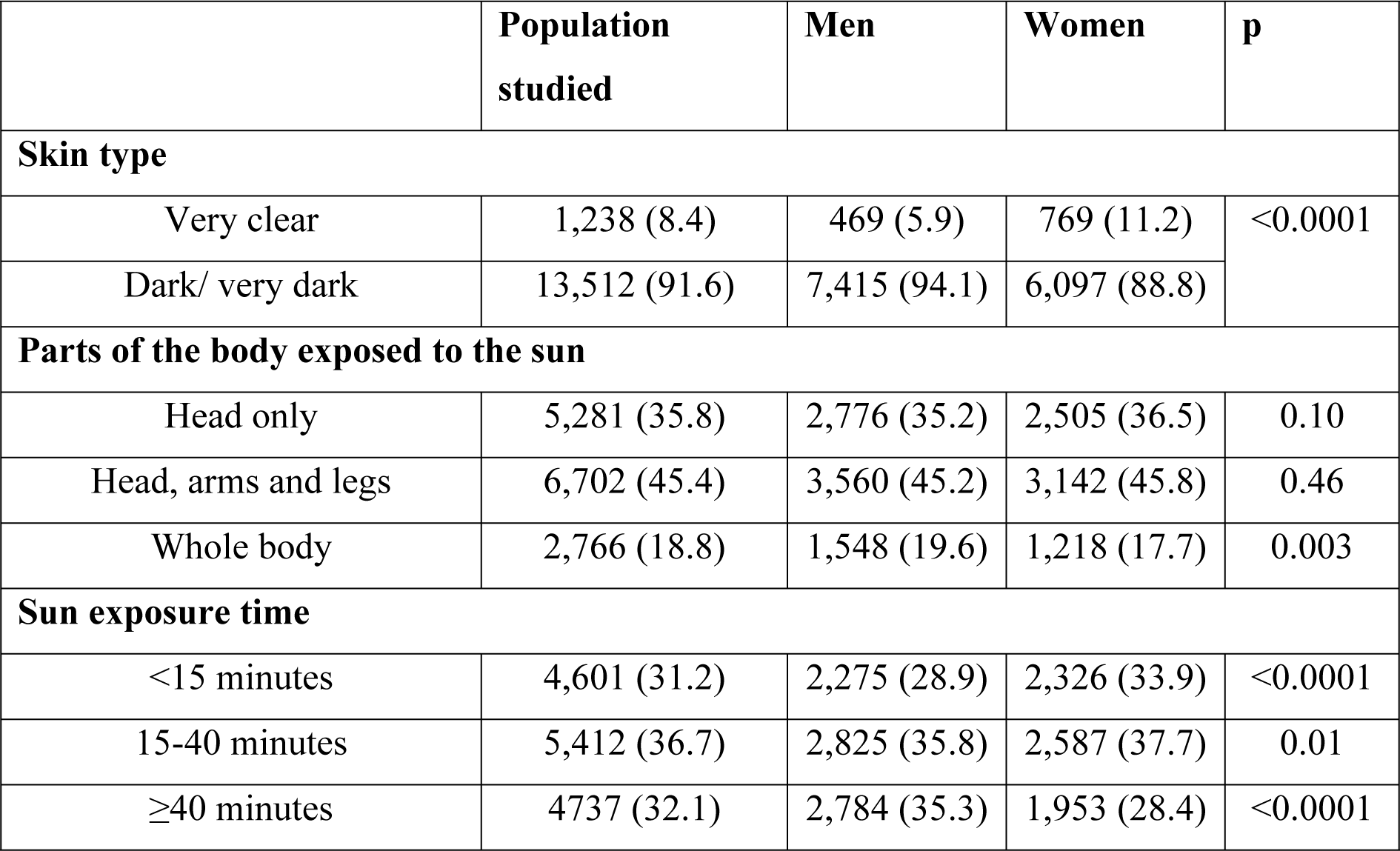
Skin type and level of sun exposure.

Still in the population studied, only 2,766 (18.8%) exposed their whole body to the sun, 1,548 (19.6%) men compared with 1,218 (17.7%) women (p=0.003).

31.2% had been exposed to the sun for less than 15 minutes.

Longer exposure > 40 minutes was more common in men than in women (35.3% vs. 28.4%; p<0.0001).

### III.3 Screening for and treatment of vitamin D deficiency

The level of Vit D screening and supplementation is shown in table 1. It shows that 2,509 (17.0%) participants had already had at least one vitamin D test and 3,593 (24.4%) were taking vitamin D supplements.

### III.4 Dietary intake of vitamin D

Dietary intakes of Vit D are shown in tables 4 and 5. The median was 2.5 (2.0-4.2) days per week of consumption of foods rich in Vit D and 5.6 (3.2-9.1)/100 gr/day of average Vit D content provided by these foods (Table 4).

**Table 4.**
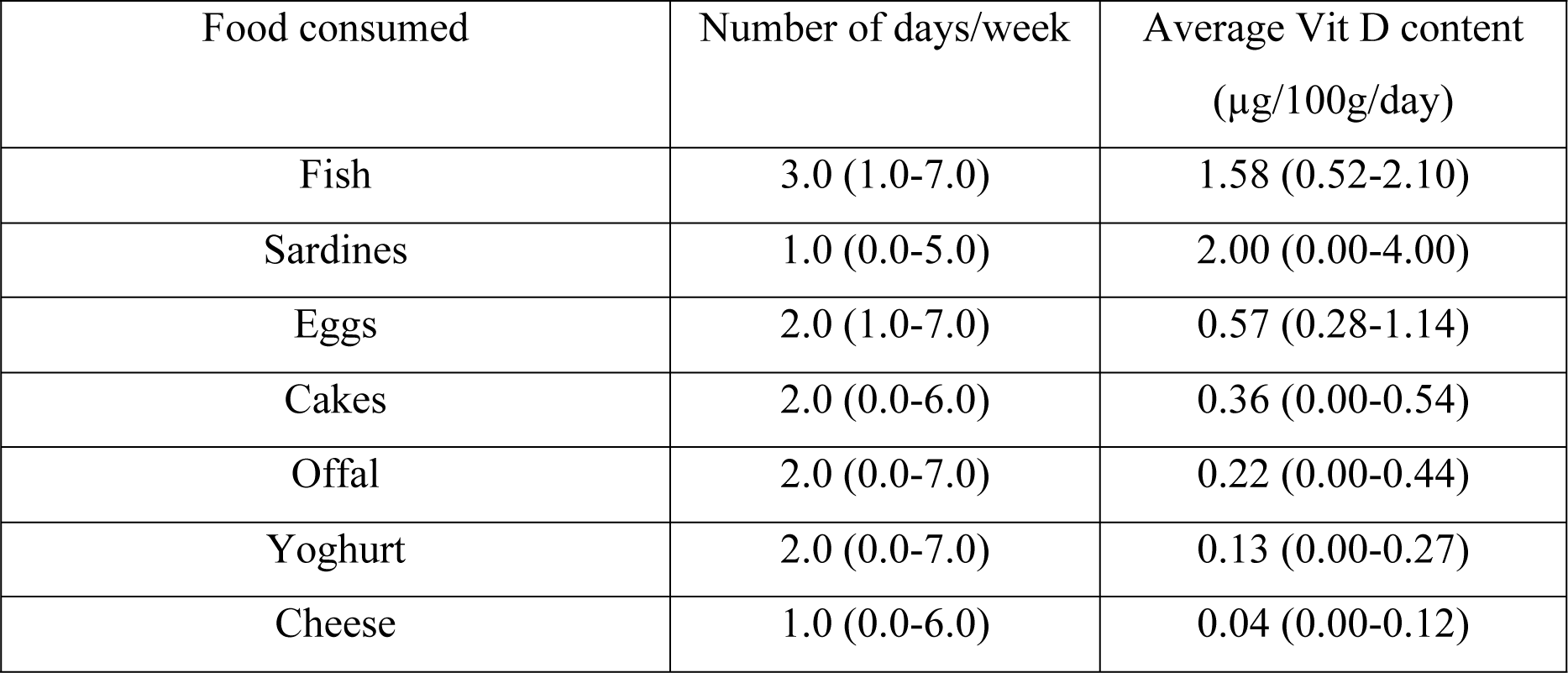
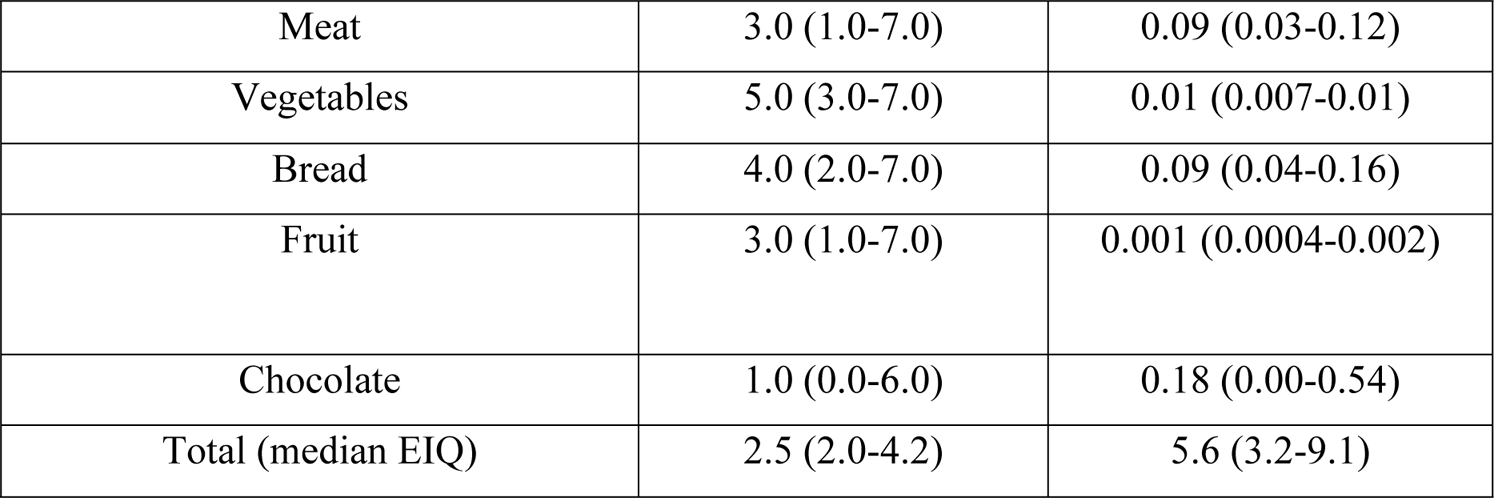
Dietary intake of vitamin D as a function of the number of days per week and the average vitamin D content.

Fish was the main source of Vit D intake in terms of frequency of consumption per week [3.0 (1.0-7.0) days/week] and average Vit D content [ 1.58 (0.52-2.10) µg/100 g/day] (Table 4).

The average Vit D content consumed per day was significantly higher in women (6.8 µg/100 gr) than in men (6.4 µg/100 gr) (p<0.0001), in participants < 40 years old (6.6 µg/100 gr) than in those between 40 and 59 years old (6.3 µg/100 gr) and > 60 years old (6.3 µg/100 gr) (p<0.0001), in the city of Lubumbashi (6.8 µg/100 gr) than in the cities of Bukavu (6.6 µg/100 gr) and Kindu (5.7 µg/100 gr) (p<0.0001) and in participants with a high socioeconomic level ( 6.7 µg/100 gr) than in those with a low socioeconomic level ( 6.5 µg/100 gr) (p=0.0009) (Table 5).

**Table 5.**
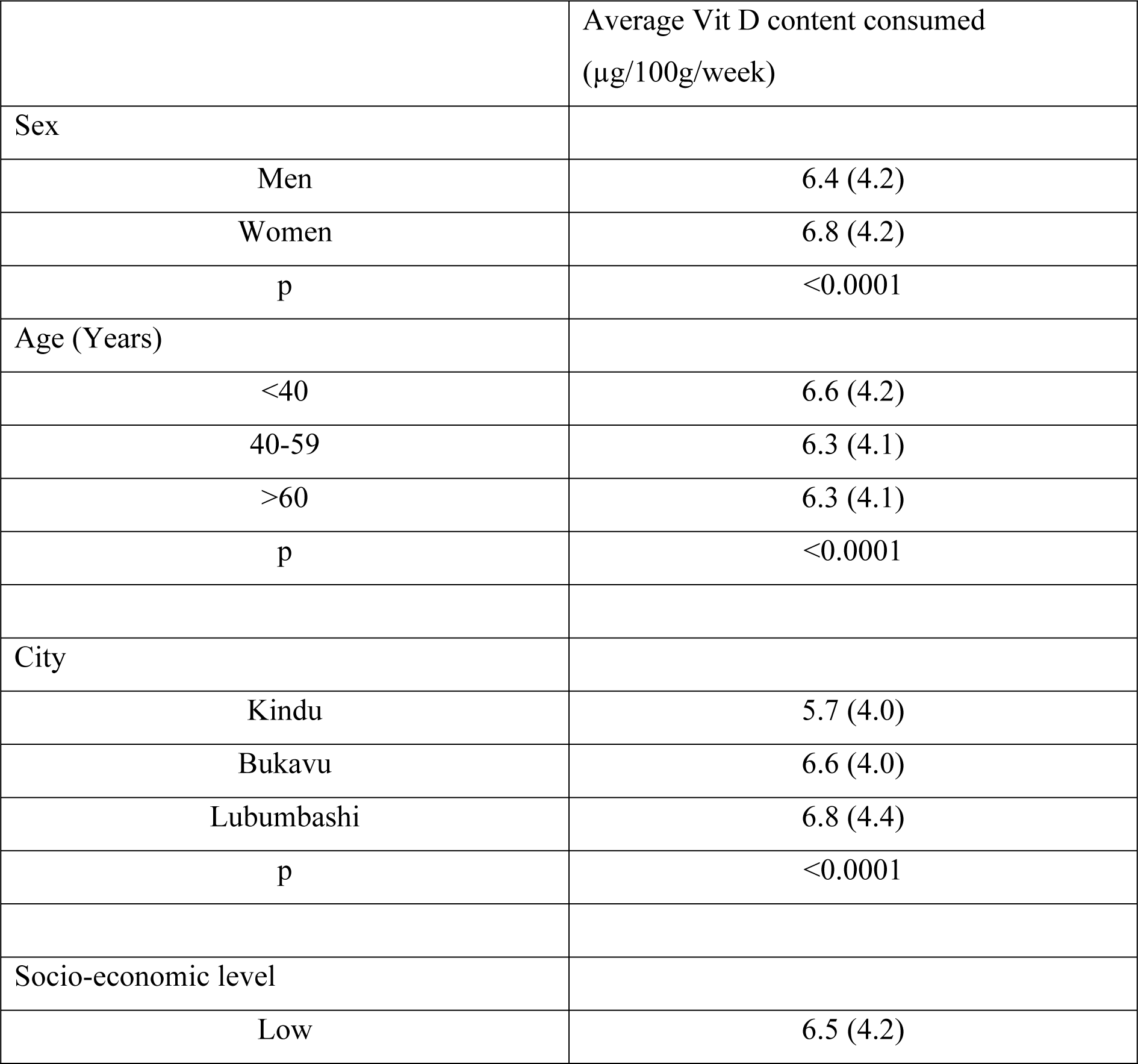
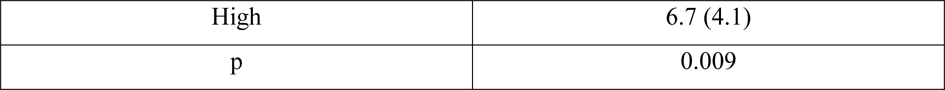
Average vitamin D intake as a function of demographic and socio-economic parameters.

### III.5 Predictors of satisfactory Vitamin D intake

Figure 1 shows the proportion of subjects with a Vit D intake judged satisfactory > 15 µg/day in this study. Only 729 (4.9%) had a satisfactory intake of Vit D. Table 6 shows the odd ratio of satisfactory Vit D intake by hypothesised factors. The results of the multivariate logistic regression show that, compared with participants aged ≥ 60 years (reference category), subjects aged 40 to 59 years had a satisfactory intake of Vit D 1.66 times more frequently (adjusted OR= 1.66; p=0.07) and those aged < 40 years a satisfactory intake 2.45 times significantly more frequently (adjusted OR= 2.45; p=0.0007).

**Figure 1.**
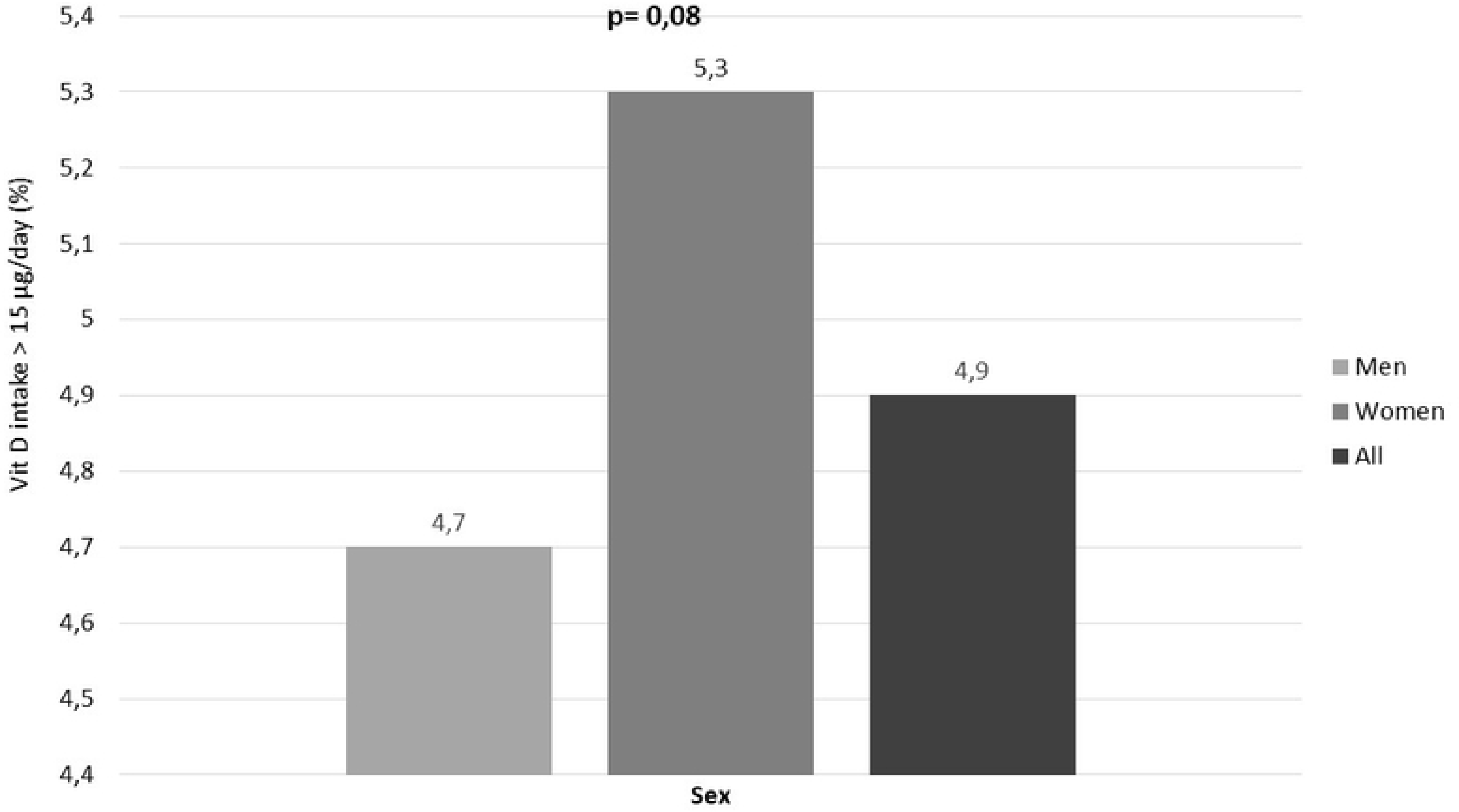
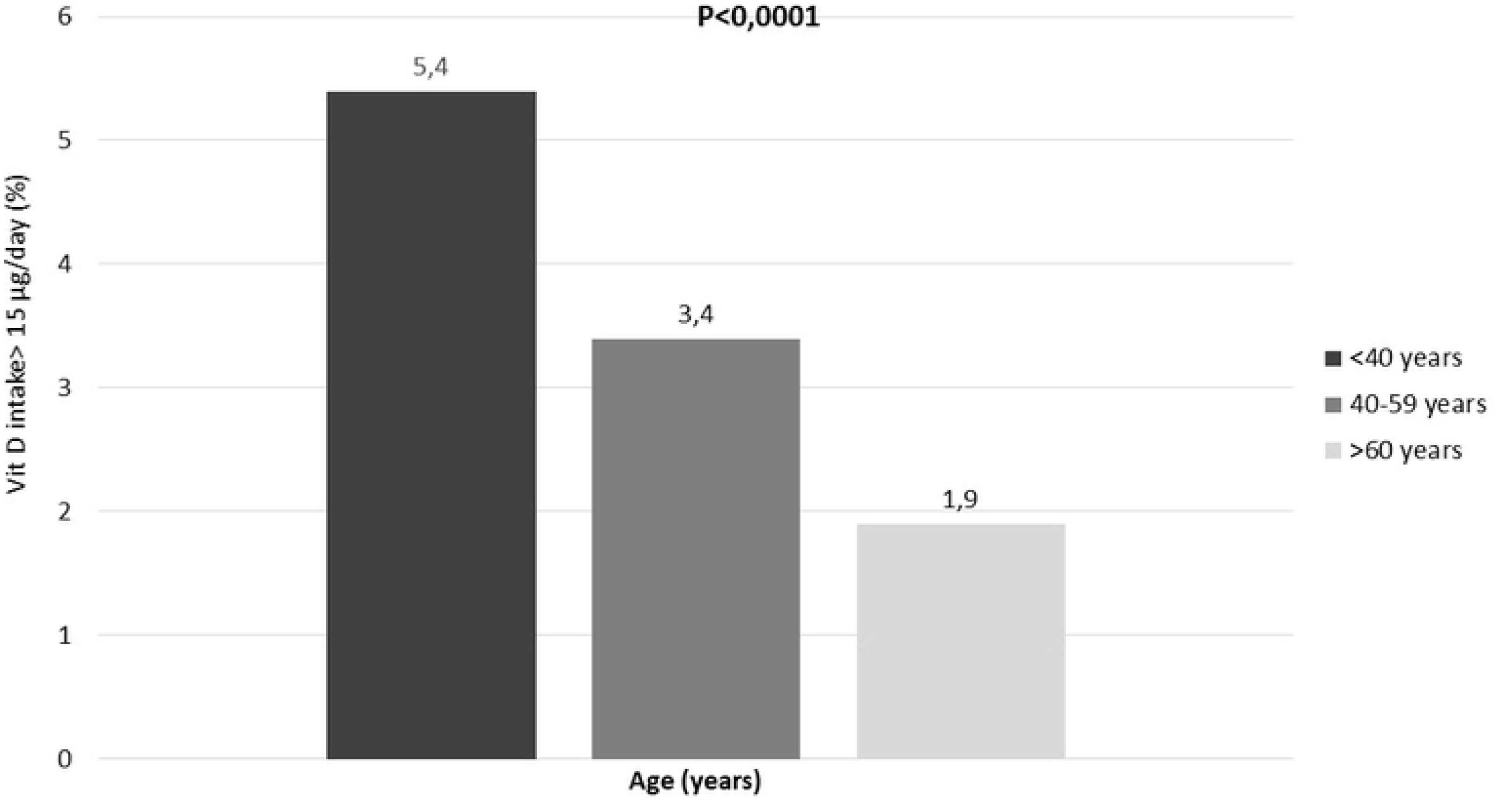
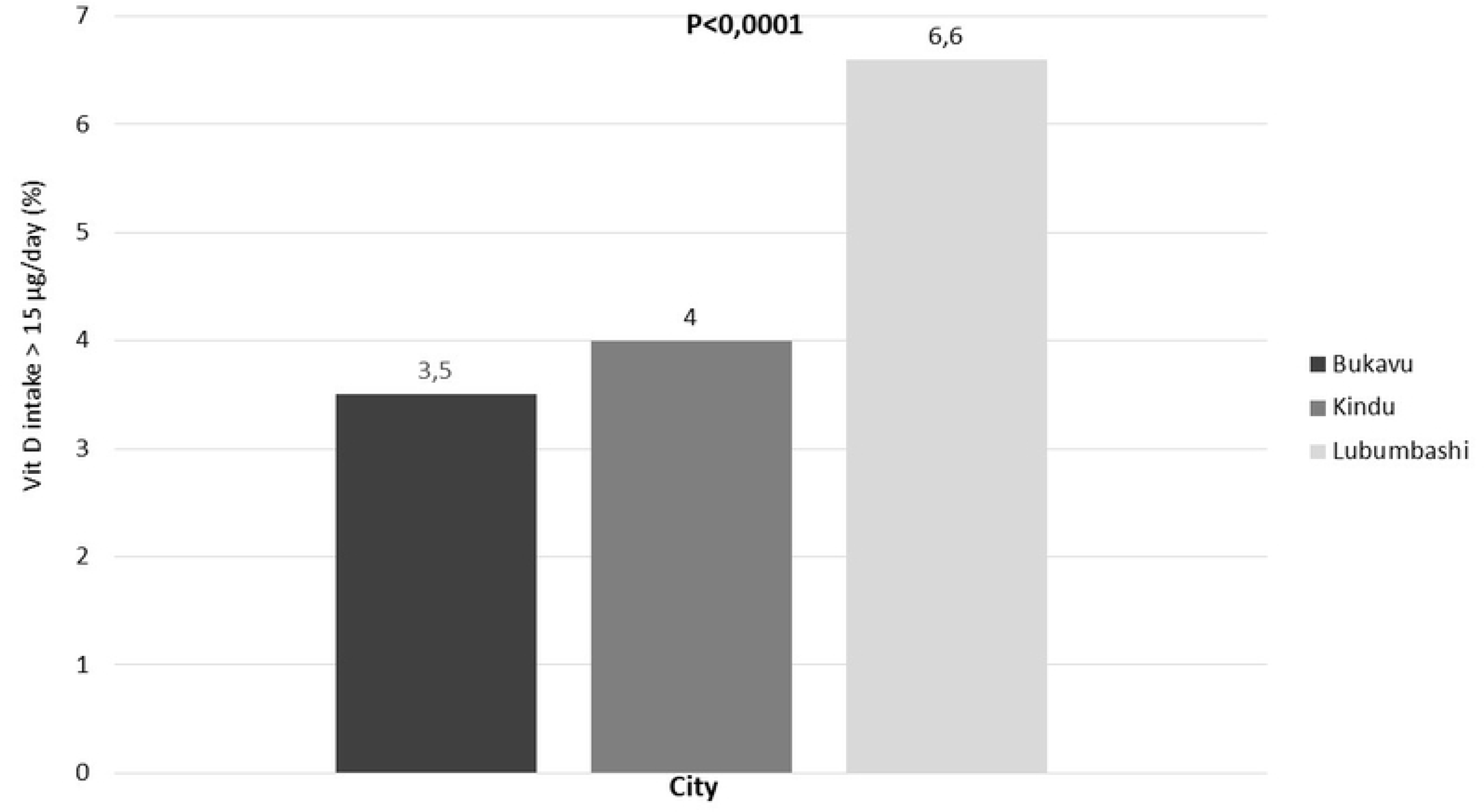
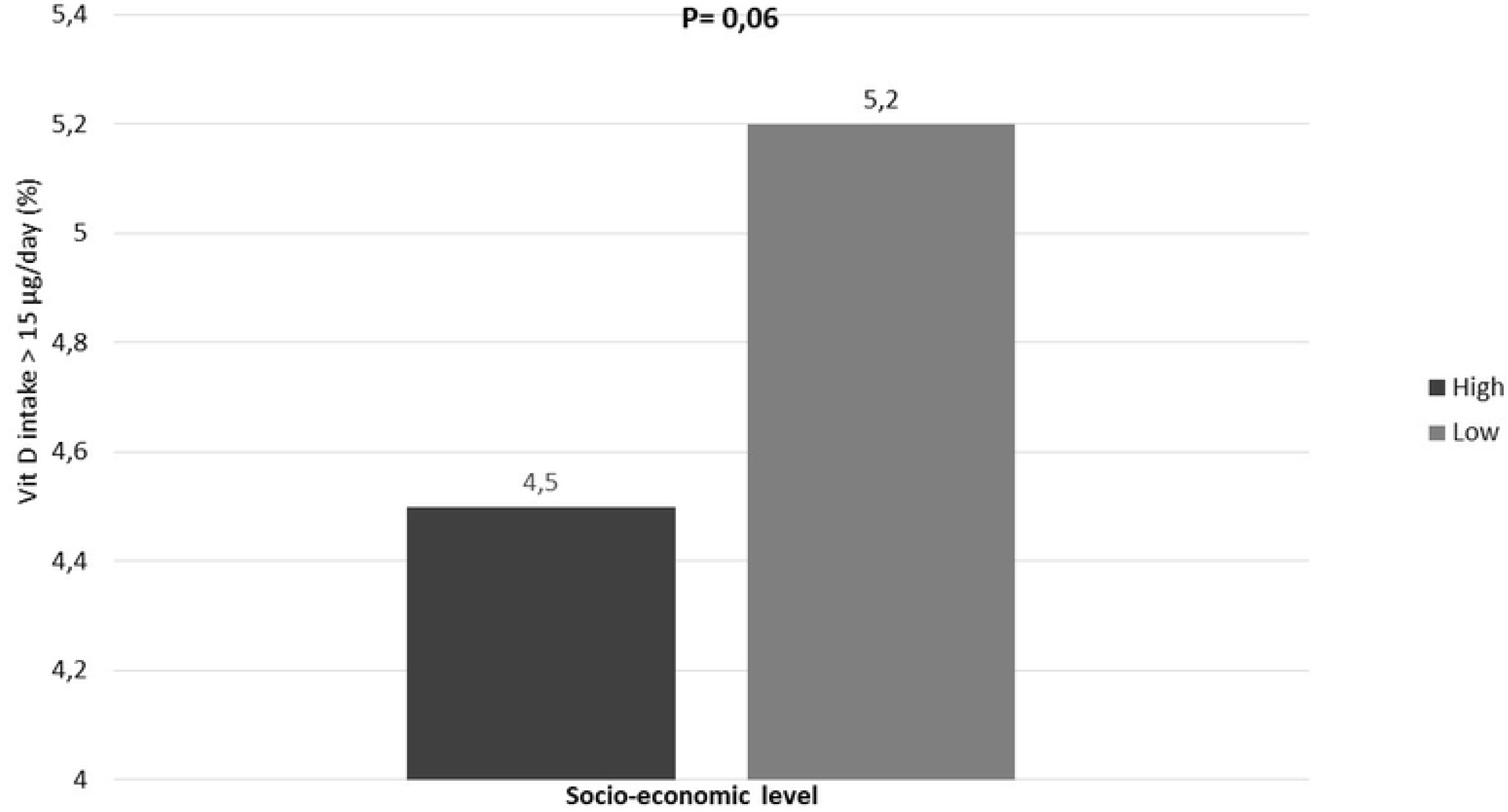
Proportion of subjects with a Vit D intake judged satisfactory > 15 µg/day: a. by gender b. by age group c. by city d. according to socio-economic level

**Table 6.** Odd ratio for a satisfactory intake of Vit D.

|  | **Crude OR**<br>**(95% CI)** | **p** | **Adjusted OR**<br>**(95% CI)** | **p** |
| --- | --- | --- | --- | --- |
| **Sex** |  |  |  |  |
| Men | 1 | - | 1 | - |
| Women | 1.14<br>(0.98-1.32) | 0.08 | 1.14<br>(0.98-1.32) | 0.08 |
| **Age (years)** |  |  |  |  |
| $<40$ | 2,90<br>(1,73-4,87) | $<0,0001$ | 2,45<br>(1,45-4,12) | 0,0007 |
| 40-59 | 1,78<br>(1,01-1,13) | 0,04 | 1,66<br>(0,94-2,92) | 0.07 |
| $>60$ | 1 | - | 1 | - |

| City |  |  |  |  |
| --- | --- | --- | --- | --- |
| Kindu | 1 |  | 1 |  |
| Bukavu | 0.87<br>(0.67-1.13) | 0.32 | 0.88<br>(0.67-1.56) | 0.37 |
| Lubumbashi | 1.67<br>(1.30-2.14) | <0.0001 | 1.64<br>(1.28-2.11) | <0.0001 |
| Socio-economic level |  |  |  |  |
| Low | 1,16<br>(0,99-1,37) | 0,06 | 1,23<br>(1,04-1,45) | 0,01 |
| High | 1 |  | 1 |  |

Compared to subjects with a high socio-economic level (reference category), subjects with a low socio-economic level had a satisfactory Vit D intake that was 1.23 times higher (adjusted OR= 1.23; p=0.01).

Finally, compared with the city of Kindu, the city of Lubumbashi was 1.64 times more likely to have subjects with a satisfactory intake of Vit D (adjusted OR= 1.64; p<0.0001).

## IV. Discussion

Studies of vitamin D intake in the general population are very rare in SSA. This study is the first to be carried out in the DRC. It was multicentre and involved a very large sample of 14,750 adult subjects.

A number of observations are worth noting.

Firstly, a large proportion of participants had very fair skin (8.4%), particularly women (11.2%). This frequency is significantly higher than that reported in the literature, which is less than 1% in SSA [7]. There are two main reasons for this: a high degree of mixed race in the population studied and the use of lightening products by women in particular. Nevertheless, >90% of the population studied had dark or very dark skin, which underlies poor cutaneous synthesis of Vit D [12]. However, several authors found no racial difference in the capacity for cutaneous synthesis of Vit D [13,14].

Secondly, the level of exposure to the sun was very low in the population studied. In fact, in this study, 31.2% of the population studied had less than 15 minutes’ exposure to the sun and 35.8% only exposed their heads to the sun. In addition, vitamin D screening was very low (17.0%). These results corroborate those of other studies which point to cultural and socio-economic factors influencing vitamin D deficiency despite the high levels of sunshine in SSA. Singularly, in the DRC, the Millennium Development Goals do not give high priority to non-communicable diseases in general or to vitamin D deficiency in particular in activities linked to national development. However, vitamin D deficiency is generally associated with other public health problems in this region, such as malaria, tuberculosis, HIV and diabetes mellitus [2,7]. Integrating a Vit D programme into already funded tuberculosis, HIV and malaria programmes would therefore make it possible to treat Vit D deficiency in the general population at a lower cost.

The third observation is a low intake of Vit D in terms of frequency of weekly consumption (2.5 days/week) and average Vit D content (5.6 µg/100 g/day) in the population studied. It should be noted that, in this study, the estimated dietary intake of Vit D was standardised using the average Vit D content of each food in µg/100 g/day proposed by Ciqual. This methodology made it possible to compare the different sub-groups and to estimate the proportion of participants with a vitamin D intake considered satisfactory (>15 µg/day). Admittedly, the methodology of this study does not allow its results to be compared with those of other studies which have attempted to estimate the daily Vit D intake in µg/day by quantifying the foods consumed [6]. However, quantification certainly has its limitations, as the foods consumed were not weighed. In our opinion, standardisation would make it possible to standardise the estimate of dietary vitamin D intake in the same way as standardisation of other formulae [15]. However, the limitation here would be that the quantity of food consumed could be over- or underestimated.

In the present study, food sources rich in Vit D were rare. Fish was the main source of Vit D. This observation could be explained by the proximity of the towns studied to streams, rivers, lakes and the Congo River. These results corroborate those of other authors [16,17].

Finally, the factors that independently influenced satisfactory daily intake of Vit D were young age < 40, region and low socio-economic level.

In relation to age, the decrease in Vit D intake with age in this study may be explained by relative restrictions in elderly people with an already high cardiovascular risk on the consumption of animal-based food (milk, eggs, meat, cheese, etc.) that are rich in Vit D but also rich in cholesterol, one of the major modifiable cardiovascular risk factors [18]. However, the results of this study are not similar to those of other authors who found either that there was no significant difference in dietary Vit D intake between different age groups [19], or that Vit D intake increased with age [20]. Nevertheless, in these studies, there was a significant increase in Vit D deficiency with age in the populations studied, which suggests that the increase in Vit D intake with age in these study populations was measures to compensate for Vit D deficiency in older subjects, especially as these regions are less sunny.

In the present study, an inverse correlation was observed between satisfactory Vit D intake and socioeconomic level. These results do not corroborate those of others who have shown that intake of vitamin and mineral supplements increased significantly with household income and level of education [20]. The inverse correlation between socioeconomic level and Vit D consumption found in our study can probably be explained by the moderation in the consumption of these foods, which are also rich in cholesterol, by participants with a high socioeconomic level compared with participants with a low socioeconomic level and less education. These results corroborate the literature. Indeed, it has been shown that risk behaviours tend to shift from higher to lower socio-economic groups as a country’s economic transition progresses. Conversely, lower socio-economic groups tend to adopt risky behaviours later in a country’s economic development [22, 23].

The results of this study must be interpreted in the light of their limitations. Firstly, the cross-sectional methodology used in this study does not allow a causal link to be established between dietary intake and serum Vit D levels. Secondly, Vit D levels were not measured in this study, which meant that it was not possible to estimate the prevalence of vitamin D deficiency in the population studied or to assess the association between this deficiency and inadequate dietary intake of Vit D.

Furthermore, as mentioned above, the standardisation of vitamin intake in µg/100g/day meant that the results of this study could not be compared objectively with those of other studies which have attempted to quantify daily dietary intakes.

## V. Conclusion

The results of the survey on Vit D intake in the general population of the DRC show a low level of exposure to sunlight and dietary intake of Vit D in this population. Integrating a Vit D programme into funded programmes such as those for tuberculosis and HIV would therefore make it possible, at a lower cost, to combat Vit D deficiency through awareness-raising, screening and treatment in the general population and in specific sub-groups.

## Data Availability

All data produced in the present work are contained in the manuscript

## Acknowledgements

This research was partially funded as part of the Seed Grant for new African Principal Investigator programme (SG-NAPI Award for 2021: SG-NAPI No. 4500454048) organised by the World Academy of Sciences (TWAS) and the German Federal Ministry of Education and Research (BMBF).

We would like to thank all those who took part in the survey for their enthusiasm.

